# Psychological status of medical students and interns in the wake of the armed conflict of April 2023 in Sudan. A cross-sectional study

**DOI:** 10.1101/2024.02.27.24303455

**Authors:** Salaheddin M. Abdulhamid, Faris Kh. Almokdad, Razan J. Q. Ahmed, Rawah M. A. M. Elbashir, Yousif Ali Ahmed Suleiman, SudaMedReCollab group

## Abstract

The ongoing civil war in Sudan of April 2023 has erupted as two belligerent groups vie for the position of ruler in the country, resulting in severe detrimental effects throughout the population, from deaths to property losses to severe deterioration of the already strained infrastructures of the country, to the sheer psychological trauma from observing and suffering the ongoing dramatic events. A subgroup is the medical students and interns in Sudan, who suffered direct losses in every regard, from sheer psychological distress to deaths, with material losses and career disruptions. We believe this is a group that should be paid due attention. This study aims to determine whether certain elements can be viewed as correlates of the psychological disorders we expect to see prevalent and their prevalences.

An online cross-sectional study was undertaken using several sampling methods to reach as far as we possibly could under the circumstances, with a structured questionnaire containing items divided between demographics, what we perceive to be factors, and scales (PHQ-9, GAD-7, AIS-8, and ITQ). The Survey was conducted from September 11^th^ to 30^th^, 2023, resulting in 7306 responses, of which 665 were excluded. A sample of 6641 were included, of which 2370 (35.7%) were males and 4271 (64.3%) were females. We found prevalences of 33.7% for depression, 22.4% for anxiety, 13.6% for PTSD, and 59.5% for Insomnia, correlating with several factors, noted to impact the psychological health of our sample. Our findings suggest the need for purposeful, directed interventions to mitigate the impact of this conflict, and all that may follow.

## Introduction

Sudan is a huge country marred with poverty and conflict, usually at the peripheries, until the civil war started in Khartoum, the capital, over who’s who in April of 2023 [1]. This has been a tragic turn of events and a deep cut to everyone, displacing approximately 9,052,822 persons according to the International Organization for Migration Displacement Tracking Matrix (IOM DTM) [2], the largest internal crisis at the time, including medical residents and medical graduates from Sudanese medical schools, which requires adaptive strategies from all stakeholders involved in the medical system and healthcare to mitigate and overcome the biopsychosocial consequences thereof.

The history of modern medical education in Sudan goes back to Kitchener School of Medicine established in 1924, currently, the School of Medicine at the University of Khartoum, and counts have risen to more than 72 schools of medicine spread across the country, with a diverse host of students who also practice as interns during and after their study, and a diversity of medical study programs [3]. This population of medical students comes from many ethnicities and backgrounds and go on to spread across the globe as practitioners of medicine in all specialties. This population is notably both vulnerable and is especially stressed at the best of times, a stress compounded in Sudan by several upheavals in recent years with deteriorating living standards that climaxed in protests regarding the price of bread despite the partial lifting of American sanctions, with an intermission involving the recent COVID-19 pandemic, and now, by an all-out war, yet this population remains poorly studied under these conditions [4].

Of the most common psychological conditions rife in healthcare workers are Depression, Anxiety, PTSD, and Insomnia due to the mental and emotional demands imposed by medical school programs, and the costs associated, be they material or otherwise, and the sheer exposure to death and disease in a developing country.

We believe this to be an important subject to study scientifically to guide appropriate intervention in a well-targeted manner as close observers ourselves who have also survived the Syrian civil war, yet suffering all the more for it, and we set out with the obvious: war will result in psychological trauma [5], and the difficulties faced will likely result in Anxiety, Depression, PTSD, and Insomnia [6–17], among other insults, for quite a while [18]. We have set out to reach our colleagues to inquire about the matter. We found a good response from which we’d included 6641 as the respondents that match the criteria.

We hypothesized that our population will be impacted by what we ourselves have noted to impact us, as well as what we have observed to impact others firsthand from our vintage points. Medical schools have suspended their role and are shut down at least partially and temporarily even as we were teaching assistants, leaving our medical students unable to complete their studies and exposing them to a myriad of risk factors for psychological trauma, also inherent to war.

The rationale is that understanding the psychological status of medical students during the current armed conflict in Sudan is crucial for addressing their mental health needs and for designing appropriate support systems in response to the current situation, as well as to others to follow. Previous research has highlighted the negative psychological consequences of armed conflicts on individuals overall [7, 16, 17, 19–22], but there is a dearth of studies focusing specifically on medical students in conflict-affected regions [23] and to our knowledge this is the first of its kind to focus on sub-Saharan Africa. Examining the psychological impact on this particular group will provide valuable insights into the experiences of medical students and interns to help develop targeted interventions to mitigate the adverse effects perhaps of both the ongoing conflict as well as others that may, unfortunately, follow, by observing the prevalence of certain psychological disorders and their correlates.

## Materials and methods

### Study design, setting, and period

An online cross-sectional survey was conducted on medical students and interns who were studying and have graduated from approximately 72 Sudanese medical schools at the beginning of the civil war, using a secure survey platform during September 11th to 30th, 2023. The data was collected anonymously while attempting to maximize participation to result in statistically significant and representative sample as far as conditions could permit. Data collectors were blinded to ensure confidentiality.

### Sample size and sampling technique

The sample size calculation was performed using the formula 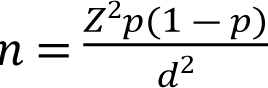 and was based on 𝑑 previously reported prevalence of depression in medical students during the COVID-19 Lockdown period in Sudan [24]. In this formula 𝑛 represents the calculated sample size, 𝑝 denotes the depression prevalence from the earlier study (0.75), 𝑍 is this chosen confidence level (1.69), and 𝑑 indicates the desired level of precision (0.01). The resulting sample size was determined to be 5355 students.

To ensure a more comprehensive representation and greater generalizability of the findings, and considering the possibility of missing responses, an additional 20% was added to the minimum required sample size, bringing it to 6426. Respondents were not pressured into answering and it was made clear that the data to be collected will be for research purposes.

Convenience, referral, and snowball sampling were the methods employed to select participants for this study considering the circumstances. Medical students were recruited based on their accessibility and willingness to participate while being encouraged to introduce others to the study. Efforts were made to reach out to a diverse range of medical students by utilizing various channels such as social media groups and batch coordinators. However, it is important to acknowledge that these methods of sampling may have introduced selection bias, as individuals who are easily accessible or motivated to participate may differ from the general population of medical students, and therefore efforts were made to minimize this bias. This was achieved by actively promoting participation among medical students from different medical schools and geographic regions affected by the armed conflict, and those who have fled Sudan altogether, with an aim at a considerable sample size. Those whose contributions were found to be significant were noted accordingly as contributors.

### Participants

Those included will be currently enrolled medical students in a Sudanese medical school, regardless of the stage of study (i.e., undergraduates), and those who are fresh postgraduate students, (i.e., interns), while excluding Individuals who were previously enrolled in a medical school but have permanently withdrawn or discontinued their studies or switched their field of study prior to the conflict, and non-medical college students.

### Study instruments

The study utilized a structured questionnaire of 49 items administered through an online survey platform across several parts. The questionnaire included validated Arabic and English versions (S1 Questionnaire English, S2 Questionnaire Arabic) as follows:

The patient health questionnaire (PHQ-9) for depression, the Generalized Anxiety Disorder Scale (GAD-7) questionnaire, the International Trauma Questionnaire (ITQ) for PTSD, and the Athens Insomnia Scale (AIS-8). The questionnaire collected information related to depression, anxiety, sleep disorders, and PTSD symptoms as the core of the study. Demographic information, such as age, gender, socioeconomic status, and proximity to conflict areas, have also been included.

The first part of the structured questionnaire is an introduction to the questionnaire and where consent is provided.

The second part of the structured questionnaire contains demographic characteristics, including age, gender, year of study, and SocioEconomic Status (SES).

The third part questionnaire aims to understand the impact of the armed conflict on individuals’ lives, Respondents were asked about their residential location’s proximity to conflict zones, their current residential status, and explores experiences of violence, changes to socioeconomic status, living arrangements, personal losses, and trauma to family members, friends, or colleagues.

The fourth part involved a patient health questionnaire known as PHQ-9 which has a sensitivity of 88% and specificity of 85%, to assess the severity of depression [25]. This validated 9-item questionnaire in both Arabic and English versions [26, 27] allows individuals to rate their depressive symptoms on a scale of 0 (not at all) to 3 (nearly every day) for each of the 9 questions [27]. The total score ranges from 0 to 27, and based on the scores, depression levels were categorized as minimal (0-4), mild (5-9), moderate (10-14), moderately severe (15-19), or severe (20-27) [28]. A PHQ-9 cut-off score of ≥15 was chosen to detect depressive symptoms, with a positive predictive value (PPV) of 51% [27].

The fifth part of the questionnaire in both Arabic and English versions involves the questionnaire for PTSD, the ITQ with its diagnostic algorithm which involves the endorsement of at least one symptom from each of the following: the Reexperience cluster, AND the Avoidance cluster, AND the sense of Current threat cluster, AND the indicators of functional impairment (due to these symptoms) [29–31].

The sixth part of the questionnaire involves the questionnaire for GAD: GAD-7. This questionnaire involves 7 items with a Sensitivity of 89%, and Specificity of 82% [32]. This questionnaire has been validated in both Arabic and English versions [26, 33, 34], and was filled with a Likert scale from 0 (Not at all), through 1 (Several days), 2 (More than half the days), to 3 (Nearly every day). the ranges are then as follow: 0-4 is considered Normal, 5-9 as Mild, 10-14 as Moderate, and 15-21 as Severe [35]. a Cut-off point of ≥15 is taken to indicate anxiety symptoms despite the loss in sensitivity, becoming 48% [32].

The seventh part of the questionnaire involves the questionnaire for sleep difficulties. The AIS-8 is a validated questionnaire [36, 37], which is composed of 8 questions, evaluating Onset of sleep, waking during the night and early morning, duration of sleep, and quality of sleep, frequency and duration of complaints, and the distress associated with insomnia, and daytime dysfunction, using a Likert scale of 0-3, with a cut-off score of 6 suggested by Soldatos et. al as they found it spots 9/10 cases [36, 38]

The eighth part of the questionnaire is for follow-up studies, requesting email address for contact at the volunteer’s discretion.

### Statistical analysis

The data collected via the questionnaire were imported into MS Excel 2016, then into IBM SPSS version 25 for analysis, and Normality was tested via the Kolmogorov-Smirnov test, finding the data to be non-Gaussian, hence the expression of data as “Median (25^th^-75^th^ quartiles)”. Mann-Whitney U-test was utilized to determine differences in PHQ9, GAD-7, and AIS-8 scores in binomial categories, and Kruskal-Wallis H-Test was used to determine differences in multinomial categories, and the Descriptives included frequencies, percentages, means, medians, and interquartile ranges considering the non-parametric nature of the data. Chi-Squared test of independence determined the associations between study variables and the psychiatric conditions in question, Spearman correlation assessed the relationship between mental health parameters, age, level of education, SES change, and number of events experienced. A binominal logistic regression model served to tease out the effect of study variables, and the likelihood of participants suffering depression, anxiety, PTSD, and insomnia. Nagelkerke R-Squared calculated the explained variation. Omnibus test of coefficients showed the overall statistical significance of the prediction model. Adjusted Odds Ratios (AOR) were expressed with their calculated 95% CI. Statistical significance was set at *p* < 0.05 level.

### Ethical approval

Ethical approval was obtained from the Quality Directorate Office – Merowe Medical City, Merowe, Northern State, Sudan.

All participants provided written consent before participating in the study, without any identification data. Emails were collected at the end of the survey at the discretion of participants for follow up studies, after completing the survey to avoid bias.

## Results

The Survey was conducted in September of 2023, resulting in 7306 responses, of which 665 were excluded, and 6641 were included (Table 1), of which 2370 (35.7%) were males and 4271 (64.3%) were females. The mean age of study participants was 22.57 ± 2.68 years, and 2801 (42.2%) students were in the preclinical years. Of the participants, 67.7% were of middle socioeconomic status, 4734 (71.3%) were displaced, and 1016 (15.3%) were in conflict zones at the time of the study. We found overall prevalences of 33.7% for depression, 22.4% for anxiety, 59.5% for Insomnia, and 13.6% for PTSD.

**Table 1.** Participant general characteristics and median (25th–75th quartile) scores of PHQ-9, GAD-7, and AIS-8 crossed with participants characteristics.

| Variables | All participant<br>n (%) | Depression score<br>Median (Q1-Q3) | Anxiety Score<br>Median (Q1-Q3) | Insomnia score<br>Median (Q1-Q3) |
| --- | --- | --- | --- | --- |
| **Overall** | n=6641 | 11 (7-17) | 8 (4-14) | 7 (4-11) |
| **Age** |  |  |  |  |
| <21 | 1522 (22.9) | 10 (6-16) | 8 (4-14) | 7 (3-11) |
| 21-24 | 3594 (54.1) | 11 (7-17) | 8 (4-14) | 7 (4-11) |
| >24 | 1525 (23) | 12 (7-18) | 8 (4-14) | 8 (4-13) |
| p-value |  | <0.001** | 0.233 | <0.001** |
| **Gender** |  |  |  |  |
| Male | 2370 (35.7) | 9 (5-16) | 7 (3-13) | 6 (3-11) |
| Female | 4271 (64.3) | 12 (8-18) | 9 (5-15) | 7 (4-12) |
| p-value |  | <0.001** | <0.001** | <0.001** |
| **Marital Status** |  |  |  |  |
| Single | 6352 (95.6) | 11 (7-17) | 8 (4-14) | 7 (4-11) |
| Married | 248 (3.7) | 12 (8-17) | 9 (5-14) | 8 (4-13) |
| others (Divorced, Widowed, etc.) | 41 (0.6) | 13 (8-19) | 9 (5-15) | 8 (4-14) |
| p-value |  | 0.126 | 0.238 | 0.006* |
| **Socioeconomic status** |  |  |  |  |
| Low-income | 1715 (25.8) | 13 (8-19) | 10 (6-16) | 8 (5-13) |
| Middle-income | 4499 (67.7) | 10 (6-16) | 8 (4-13) | 7 (3-11) |
| High-income | 427 (6.4) | 8 (4-15) | 6 (2-12) | 5 (2-10) |
| p-value |  | <0.001** | <0.001** | <0.001** |
| **Level of Education** |  |  |  |  |
| Pre-clinical year | 2801 (42.2) | 11 (7-17) | 8 (4-14) | 7 (3-11) |
| Clinical year | 2349 (35.4) | 11 (7-17) | 8 (4-14) | 7 (4-11) |
| Internship | 1491 (22.5) | 11 (7-18) | 8 (5-14) | 8 (4-13) |
| p-value |  | 0.005* | 0.345 | <0.001** |
| **Living arrangement** |  |  |  |  |
| With family | 5957 (89.7) | 11 (7-17) | 8 (4-14) | 7 (4-11) |
| With friends and colleagues | 498 (7.5) | 12 (8-18) | 9 (6-15) | 8 (5-13) |
| Alone | 186 (2.8) | 13 (8-20) | 11 (6-16) | 9 (4-14) |
| p-value |  | <0.001** | <0.001** | <0.001** |
| **Internal\external displacement** |  |  |  |  |
| Non-displaced | 1907 (28.7) | 10 (6-16) | 7 (3-13) | 6 (3-9) |
| Internally displaced | 1690 (25.4) | 11 (7-17) | 9 (5-14) | 7 (4-11) |
| Externally displaced | 3044 (45.8) | 12 (7-18) | 9 (5-14) | 8 (4-12) |
| p-value |  | <0.001** | <0.001** | <0.001** |
| **Living in conflict zone** |  |  |  |  |
| Yes | 1016 (15.3) | 12 (8-18) | 9 (5-15) | 7 (4-12) |
| No | 5625 (84.7) | 11 (7-17) | 8 (4-14) | 7 (4-11) |
| p-value |  | 0.005* | <0.001** | 0.004* |

\*Significant at p <0.05, \*\*Significant at p <0.001.

The total respondents suffering loss of family members, friends, or colleagues due to the armed conflict in Sudan were 1926 (29%) (S3 Table 1). In a similar vein, 2770 (41.7%) have suffered injuries to family members, friends, or colleagues due to the armed conflict in Sudan, and 1168 (17.6%) have witnessed or suffered direct attacks, such physical assaults, bombings, shootings, kidnap, or torture, and 1990 participants (30%) have suffered destruction or loss of property, and 298 (4.5%) have suffered or witnessed gender-specific violence (GSV) (other than sexual violence, e.g. hair shaved, insults, etc…), and 72 (1.1%) have suffered or witnessed rape or sexual violence, and 1170 (17.6%) have witnessed violence against others.

Depression symptoms were assessed with the PHQ-9 scale, and the results were classified into five degrees in term of the severity of depression (Table 2). The PHQ-9 score ranged from 0 to 27, with a median of 11 (IQR: 7-17), while the mean (±SD) score of depression symptoms was 11.93 (±6.86), thus indicating moderate depression. A total of 5634 (84.8) of the participants had different degrees of depression. In addition, 2241 (33.7%) students had a PHQ-9 score of ≥15, which is indicative of moderately severe to severe depression. Suicidal ideation appeared in 2213 (33.3%) of students, as follows: several days 1203 (18.1%), more than half of the days 410 (6.2%), and 600 (9%) nearly every day.

**Table 2.** Scores, Frequencies, and Gradings using relevant measures.

| Category | Grade (score range) | Frequency | percentage |
| --- | --- | --- | --- |
| **Depression (PHQ-9)** | Minimal (0-4) | 1007 | 15.2 |
|  | Mild (5-9) | 1875 | 28.2 |
|  | Moderate (10-14) | 1518 | 22.9 |
|  | Moderately severe (15-19) | 1198 | 18 |
|  | Severe (20-27) | 1043 | 15.7 |
| **Anxiety (GAD-7)** | Normal (0-4) | 1733 | 26.1 |
|  | Mild (5-9) | 2086 | 31.4 |
|  | Moderate (10-14) | 1334 | 20.1 |
|  | Severe (15-21) | 1488 | 22.4 |
| **Insomnia (AIS-8)** | None (0-5) | 2689 | 40.5 |
|  | Mild (6-9) | 1790 | 27 |
|  | Moderate (10-15) | 1556 | 23.4 |
| | Severe ($\geq 16$) | 606 | 9.1 |
| **PTSD (ITQ)** | Avoidance | 2651 | 39.9 |
|  | Reexperience | 2651 | 33.9 |
|  | Current threat | 3026 | 45.6 |
|  | Functional impairment | 3670 | 55.3 |
|  | Met criteria for PTSD | 901 | 13.6 |

\*Significant at $p < 0.05$ , \*\*Significant at $p < 0.001$ .

The Analysis reveals a significant association between depressive symptoms and several demographic and situational variables as illustrated in (Table 3). Notably, individuals aged 21-24 exhibit a higher prevalence of depressive symptoms, and females are more susceptible than males. Socioeconomic status demonstrates a strong link, especially in those of lower income. Living alone and external displacement are associated with higher depressive symptom rates. Additionally, being in the conflict zone and exposure to injuries are linked to increased depressive symptoms, especially exposure to death and material losses. Direct attacks, destruction/loss of property, gender-specific violence, rape, or sexual violence were all linked to elevated depressive symptoms.

**Table 3.**
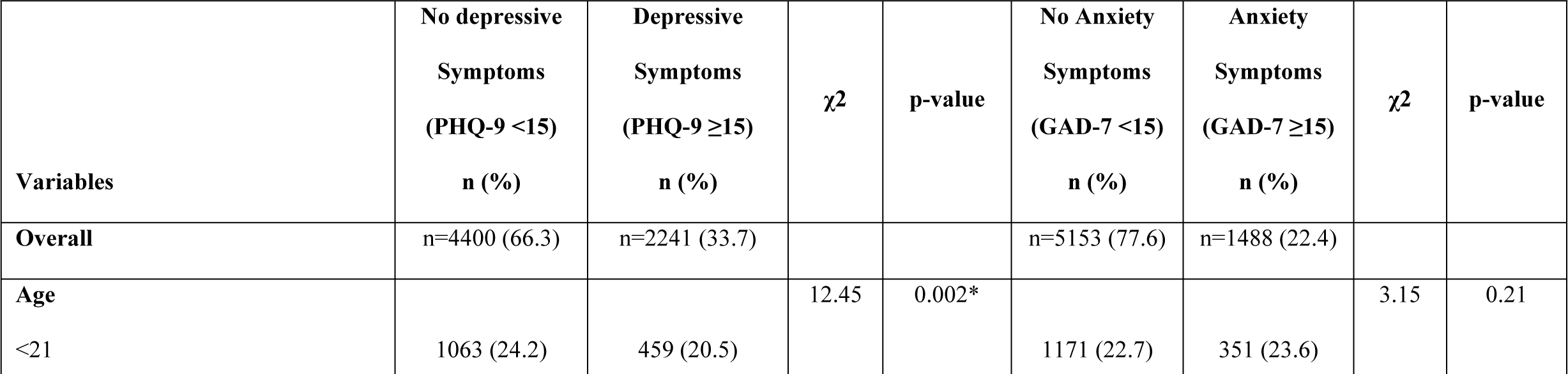
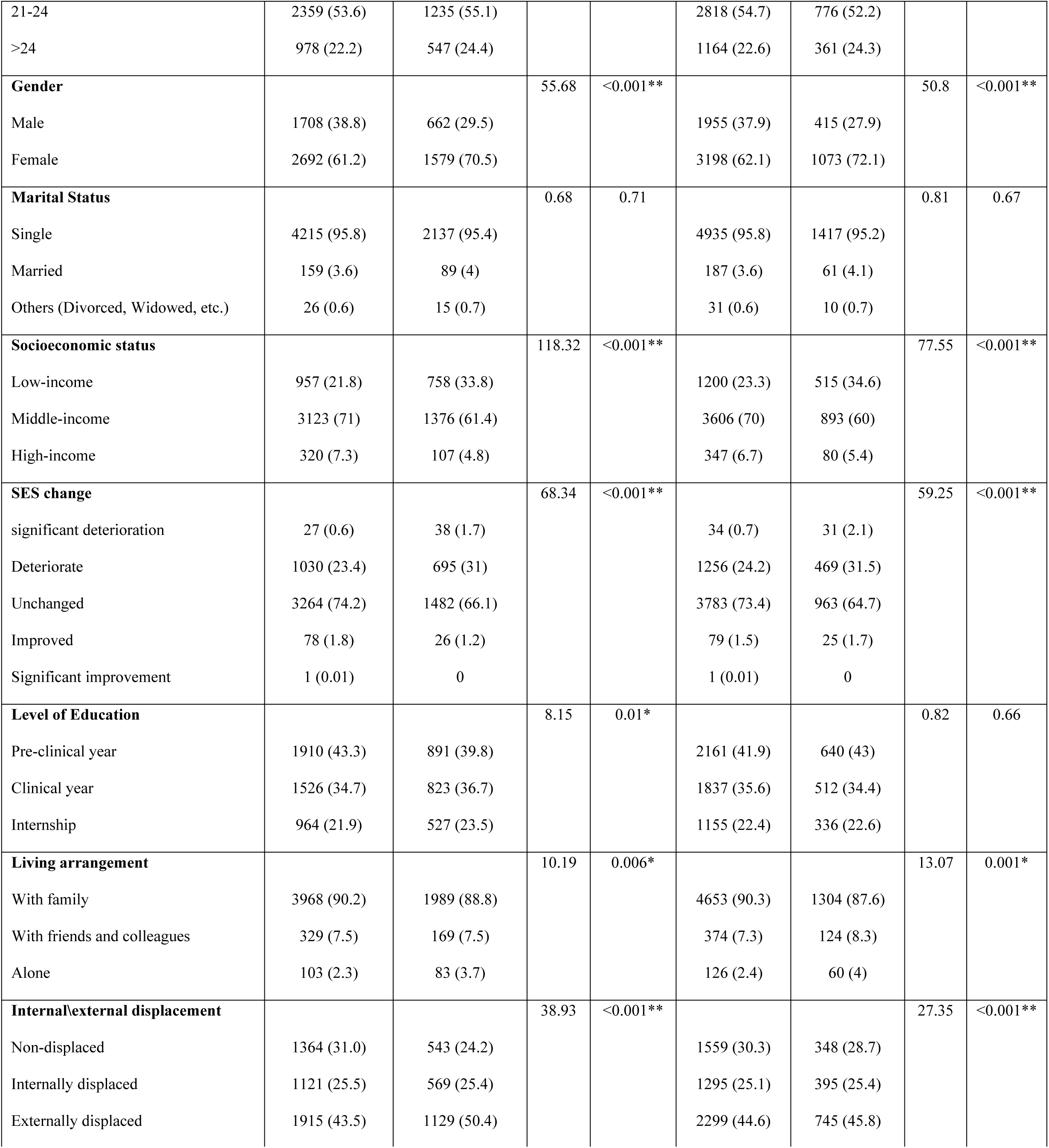
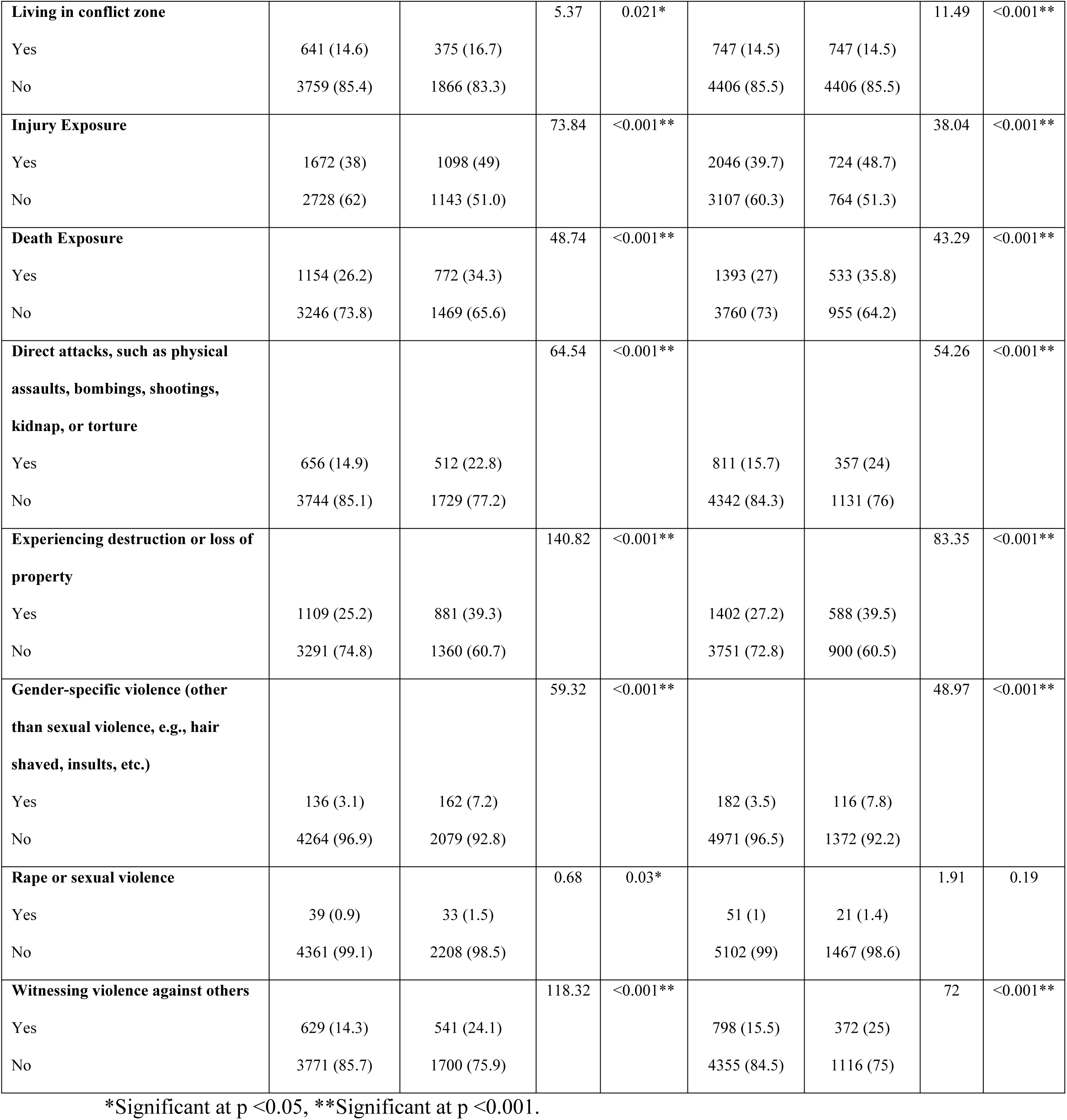
Prevalence and association of Depression and Anxiety symptoms with the study variables.

The logistical regression model for depression (Table 4) was statistically significant for ꭕ2(17) = 457.968, p<0.001, and it explained 9.2% (Nagelkerke R2) of the variance, classifying 68.21% of cases, at a sensitivity of 25% and specificity of 90%, and with a positive predictive value 56% of and negative predictive value of 70%. The most significant, impactful predictive factors are the following, in descending order: Low SES (OR: 2.06), being female (OR: 1.86), Gender-Specific Violence (OR:1.59), experiencing destruction or loss of property (OR: 1.54), living alone (OR: 1.4), witnessing violence against others (OR: 1.34), suffering direct assault and exposure to injuries (OR: 1.28), displacement (OR: 1.22), and exposure to death (OR: 1.16).

**Table 4.** Binomial logistical regression model predicting the likelihood for Depression and Anxiety symptoms based on the study characteristics.

|  | Dependent variable: Depression |  |  |  | Dependent variable: Anxiety |  |  |  |
| --- | --- | --- | --- | --- | --- | --- | --- | --- |
| Independent Variables | AOR | 95% CI of AOR |  | p-value | AOR | 95% CI of AOR |  | p-value |
|  |  | Lower | Upper |  |  | Lower | Upper |  |
| Age | 1.02 | 1.01 | 1.05 | 0.04* | 1.03 | 1.01 | 1.06 | 0.05 |
| **Gender** |  |  |  |  |  |  |  |  |
| Male | Reference |  |  |  | Reference |  |  |  |
| Female | 1.86 | 1.65 | 2.1 | <0.001** | 1.96 | 1.71 | 2.24 | <0.001** |
| **Socioeconomic status** |  |  |  |  |  |  |  |  |
| Low-income | 2.06 | 1.61 | 2.64 | <0.001** | 1.57 | 1.19 | 2.07 | 0.001* |
| Middle-income | 1.24 | 0.98 | 1.57 | 0.63 | 1.01 | 0.78 | 1.38 | 0.94 |
| High-income | Reference |  |  |  | Reference |  |  |  |
| **Level of Education** |  |  |  |  |  |  |  |  |
| Pre-clinical year | Reference |  |  |  | Reference |  |  |  |
| Clinical year | 1.08 | 0.94 | 1.24 | 0.25 | 0.86 | 0.72 | 1.01 | 0.06 |
| Internship | 0.93 | 0.77 | 1.11 | 0.44 | 0.75 | 0.61 | 0.92 | 0.007* |
| **Living arrangement** |  |  |  |  |  |  |  |  |
| With family | Reference |  |  |  | Reference |  |  |  |
| With friends and colleagues | 0.96 | 0.78 | 1.18 | 0.71 | 1.18 | 0.94 | 1.49 | 0.13 |
| Alone | 1.4 | 1.02 | 1.91 | 0.03* | 1.51 | 1.08 | 2.10 | 0.01* |
| **Displacement** |  |  |  |  |  |  |  |  |
| Non-displaced | Reference |  |  |  | Reference |  |  |  |
| Displaced | 1.22 | 1.08 | 1.39 | 0.002* | 1.30 | 1.12 | 1.51 | <0.001** |
| **Living in conflict zone** |  |  |  |  |  |  |  |  |
| No | Reference |  |  |  | Reference |  |  |  |
| Yes | 0.98 | 0.84 | 1.15 | 0.88 | 1.14 | 0.97 | 1.35 | 0.11 |
| **Injury Exposure** |  |  |  |  |  |  |  |  |
| No | Reference |  |  |  | Reference |  |  |  |
| Yes | 1.28 | 1.13 | 1.45 | <0.001* | 1.12 | 0.97 | 1.92 | 0.95 |
| **Death Exposure** |  |  |  |  |  |  |  |  |
| No | Reference |  |  |  | Reference |  |  |  |
| Yes | 1.16 | 1.01 | 1.33 | 0.02* | 1.27 | 1.1 | 1.48 | 0.001* |
| **Direct attacks, such as physical assaults, bombings, shootings, kidnap, or torture** |  |  |  |  |  |  |  |  |
| No | Reference |  |  |  | Reference |  |  |  |
| Yes | 1.28 | 1.11 | 1.48 | <0.001** | 1.29 | 1.11 | 1.51 | 0.001* |
| **Experiencing destruction or loss of property** |  |  |  |  |  |  |  |  |
| No | Reference |  |  |  | Reference |  |  |  |
| Yes | 1.54 | 1.36 | 1.74 | <0.001** | 1.41 | 1.23 | 1.61 | <0.001** |
| **Gender-specific violence** |  |  |  |  |  |  |  |  |
| No | Reference |  |  |  | Reference |  |  |  |
| Yes | 1.59 | 1.22 | 2.07 | <0.001** | 1.58 | 1.21 | 2.08 | <0.001** |
| **Rape or sexual violence** |  |  |  |  |  |  |  |  |
| No | Reference |  |  |  | Reference |  |  |  |
| Yes | 0.71 | 0.43 | 1.19 | 0.2 | 0.59 | 0.33 | 1.04 | 0.07 |
| **Witnessing violence against others** |  |  |  |  |  |  |  |  |
| No | Reference |  |  |  | Reference |  |  |  |
| Yes | 1.34 | 1.16 | 1.55 | <0.001** | 1.31 | 1.11 | 1.54 | <0.001** |

\*Significant at $p < 0.05$ , \*\*Significant at $p < 0.001$ .

Anxiety symptoms were assessed with the GAD-7 scale, and the results were classified accordingly into degrees of anxiety (Table 2). The median was 8 for mild anxiety (4-14), with a mean of 9.26 (±6.08). A total of 1488 (22.4%) scored ≥15 for severe anxiety. According to GAD-7 categorization, 1733 (26.1) of medical students demonstrate a lack of anxiety symptoms. In contrast, 4908 (73.9%) of students exhibited varying levels of anxiety: 31.4% reported mild anxiety, 20.1% moderate anxiety, and 22.4% experienced severe anxiety. The prevalence of anxiety symptoms and their association with various factors were investigated (Table 4). The logistical regression model for Anxiety (Table 4) was statistically significant for ꭕ2(17) = 334.4, p<0.001, and it explained 7% (Nagelkerke R2) of the variance, classifying 75.7% of cases, at a sensitivity of 18.9% and specificity of 92.1%, and with a positive predictive value 40.7% of and negative predictive value of 79.7%. There is a significant association across several variables. a significant gender difference emerges, with females exhibiting a higher prevalence of anxiety symptoms. Socioeconomic status demonstrates a strong association, indicating a higher prevalence among low-income individuals. injury exposure and death exposure are significantly linked to a higher prevalence of anxiety symptoms. Additionally, being exposed to direct attack, destruction/loss of property, and gender-specific violence, were all linked to increased anxiety symptoms. The most significant, impactful predictive factors are the following, in descending order: Being female (OR: 1.96), Gender Specific Violence (OR: 1.58), low SES (OR: 1.57), Living alone (OR: 1.51), loss/destruction of property (OR: 1.41), witnessing violence against others (OR: 1.31), suffering direct assault (OR: 1.29), exposure to death (OR: 1.27).

Insomnia symptoms were assessed with the AIS-8 scale, and the scores were stratified accordingly into degrees of insomnia (Table 2). In total (Table 1), the median was 7 for minimal insomnia (4-11), with a mean of 7.75 (±5.25). A total 3952 (59.5%) of participants have different degree of insomnia: as follow (33.7%) experienced mild degrees of insomnia, (10.3%) moderate, and (1.3%) severe insomnia. In term of association, statically significant associations were found in all study variables except for age group, marital status, and level of education, as showed in Table 5.

**Table 5.** Prevalence and association of insomnia and PTSD symptoms crossed with the study variables.

| Variables | No Insomnia<br>Symptoms<br>(AIS-8 <6)<br>n (%) | Insomnia<br>Symptoms<br>(AIS-8 ≥6)<br>n (%) | $\chi^2$ | p-value | No PTSD<br>Symptoms<br>Criteria not<br>met<br>n (%) | PTSD<br>Symptoms<br>Met criteria<br>n (%) | $\chi^2$ | p-value |
| --- | --- | --- | --- | --- | --- | --- | --- | --- |
| **Overall** | n=2686 (40.5) | n=3952 (59.5) |  |  | n=5740 (86.4) | n=901 (13.6) |  |  |
| **Age** |  |  | 12.51 | 0.002* |  |  | 7.76 | 0.02* |
| <21 | 51 (24.2) | 871 (22) |  |  | 1331 (23.2) | 191 (21.2) |  |  |
| 21-24 | 1477 (54.9) | 2117 (53.6) |  |  | 3123 (54.4) | 471 (52.3) |  |  |
| >24 | 561 (20.9) | 964 (24.4) |  |  | 1286 (22.4) | 239 (26.5) |  |  |
| **Gender** |  |  | 31.97 | <0.001** |  |  | 39.05 | <0.001** |
| Male | 1068 (39.7) | 1302 (32.9) |  |  | 2130 (37.1) | 238 (26.4) |  |  |
| Female | 1621 (60.3) | 2650 (67.1) |  |  | 3608 (62.9) | 663 (73.6) |  |  |
| **Marital Status** |  |  | 8.16 | 0.017* |  |  | 8.91 | 0.012* |
| Single | 2595 (96.5) | 3757 (95.1) |  |  | 5507 (95.9) | 845 (93.8) |  |  |
| Married | 82 (3) | 166 (4.2) |  |  | 199 (3.5) | 49 (5.4) |  |  |
| Others (Divorced, Widowed, etc.) | 12 (0.4) | 29 (0.7) |  |  | 34 (0.6) | 7 (0.8) |  |  |
| **Socioeconomic status** |  |  | 108.73 | <0.001** |  |  | 49.07 | <0.001** |
| Low-income | 529 (19.7) | 1186 (30) |  |  | 1397 (24.3) | 318 (35.3) |  |  |
| Middle-income | 1930 (71.8) | 2569 (65) |  |  | 3963 (69) | 536 (59.5) |  |  |
| High-income | 230 (8.6) | 197 (5) |  |  | 380 (6.6) | 47 (5.2) |  |  |
| **SES change** |  |  | 65.21 | <0.001** |  |  | 55.19 | <0.001** |
| significant deterioration | 8 (0.3) | 57 (1.4) |  |  | 55 (1) | 10 (1.1) |  |  |
| Deteriorate | 590 (21.9) | 1135 (28.7) |  |  | 1404 (24.5) | 321 (35.6) |  |  |
| Unchanged | 2051 (76.3) | 2695 (68.2) |  |  | 4195 (73.1) | 551 (61.2) |  |  |
| Improved | 40 (1.5) | 64 (1.6) |  |  | 85 (1.5) | 19 (2.1) |  |  |
| Significant improvement | 0 | 1 (0.02) |  |  | 1 (0.01) | 0 |  |  |
| **Level of Education** |  |  | 17.94 | <0.001** |  |  | 0.474 | 0.79 |
| Pre-clinical year | 1189 (44.2) | 1612 (40.8) |  |  | 2430 (42.3) | 371 (41.2) |  |  |
| Clinical year | 965 (35.9) | 1384 (35) |  |  | 2027 (35.3) | 322 (35.7) |  |  |
| Internship | 535 (19.9) | 956 (24.2) |  |  | 1283 (22.4) | 208 (23.1) |  |  |
| **Living arrangement** |  |  | 41.84 | <0.001** |  |  | 6.95 | 0.03* |
| With family | 2490 (92.6) | 3467 (87.7) |  |  | 5171 (90.1) | 786 (87.2) |  |  |
| With friends and colleagues | 140 (5.2) | 358 (9.1) |  |  | 413 (7.2) | 85 (9.4) |  |  |
| Alone | 59 (2.2) | 127 (3.2) |  |  | 156 (2.7) | 30 (3.3) |  |  |
| **Internal/external displacement** |  |  | 92.43 | <0.001** |  |  | 6.48 | 0.04* |
| Non-displaced | 923 (34.3) | 984 (24.9) |  |  | 1680 (29.3) | 227 (25.2) |  |  |
| Internally displaced | 708 (26.3) | 982 (24.8) |  |  | 1454 (25.3) | 236 (26.2) |  |  |
| Externally displaced | 1058 (39.3) | 1986 (50.3) |  |  | 2606 (45.4) | 438 (48.6) |  |  |
| **Living in conflict zone** |  |  | 18.77 | <0.001** |  |  | 10.89 | 0.001* |
| Yes | 349 (13) | 667 (16.9) |  |  | 845 (14.7) | 171 (19) |  |  |
| No | 2340 (87) | 3285 (83.1) |  |  | 4895 (85.3) | 730 (81) |  |  |
| **Injury Exposure** |  |  | 101.37 | <0.001** |  |  | 23.84 | <0.001** |
| Yes | 923 (34.3) | 1847 (46.7) |  |  | 2327 (40.5) | 443 (49.2) |  |  |
| No | 1766 (65.7) | 2105 (53.3) |  |  | 3413 (59.5) | 458 (50.8) |  |  |
| **Death Exposure** |  |  | 80.49 | <0.001** |  |  | 46.87 | <0.001** |
| Yes | 617 (22.9) | 1309 (33.1) |  |  | 1578 (27.5) | 348 (38.6) |  |  |
| No | 2072 (77.1) | 2643 (66.9) |  |  | 4162 (72.5) | 553 (61.4) |  |  |
| **Direct attacks, such as physical assaults, bombings, shootings, kidnap, or torture** |  |  | 70.56 | <0.001** |  |  | 47.9 | <0.001** |
| Yes | 345 (12.8) | 823 (20.8) |  |  | 936 (16.3) | 232 (25.7) |  |  |
| No | 2344 (87.2) | 3129 (79.2) |  |  | 4804 (83.7) | 669 (74.3) |  |  |
| **Experiencing destruction or loss of property** |  |  | 137.35 | <0.001** |  |  | 61.2 | <0.001** |
| Yes | 591 (22) | 1399 (35.4) |  |  | 1620 (28.2) | 370 (41.1) |  |  |
| No | 2098 (78) | 2553 (64.6) |  |  | 4120 (71.8) | 531 (58.9) |  |  |
| **Gender-specific violence** |  |  | 41.98 | <0.001** |  |  | 7.26 | 0.009 |
| Yes | 67 (2.5) | 231 (5.8) |  |  | 242 (4.2) | 56 (6.2) |  |  |
| No | 2622 (97.5) | 3721 (94.2) |  |  | 5498 (95.8) | 845 (93.8) |  |  |
| **Rape or sexual violence** |  |  | 11.67 | <0.001** |  |  | 10.2 | 0.003 |
| Yes | 15 (0.6) | 57 (1.4) |  |  | 53 (0.9) | 19 (2.1) |  |  |
| No | 2674 (99.4) | 3895 (98.6) |  |  | 5687 (99.1) | 882 (97.9) |  |  |
| **Witnessing violence against others** |  |  | 128.48 | <0.001** |  |  | 70.49 | <0.001** |
| Yes | 301 (11.2) | 869 (22) |  |  | 922 (16.1) | 248 (27.5) |  |  |
| No | 2388 (88.8) | 3083 (78) |  |  | 4818 (83.9) | 653 (72.5) |  |  |

\*Significant at $p < 0.05$ , \*\*Significant at $p < 0.001$ .

Females showed a higher prevalence of insomnia than males. SES significantly influence insomnia, with higher rates in low and middle income compared to high income individuals. Exposure to traumatic events further delineated the complexity of the relationship between insomnia and trauma, as individuals reported direct attack, including physical assaults, shooting, bombings, kidnapping or torture, demonstrate markedly elevated prevalence of insomnia. Similarly, those who experienced destruction or loss of property, or witnessed violence against others showed increased rates of insomnia symptoms.

The logistical regression model for Insomnia (Table 6) was statistically significant for ꭕ2(17) = 536.1, p<0.001, and it explained 10.5% Nagelkerke R2 of the variance, classifying 63.5% of cases, at a specificity of 81.3% and sensitivity of 37%, and with a positive predictive value 65.5% of and negative predictive value of 57.5%.

**Table 6.** Binomial logistical regression model predicting the likelihood for Insomnia and PTSD symptoms based on the study characteristics.

| Independent Variables | Dependent variable: Insomnia |  |  |  | Dependent variable: PTSD |  |  |  |
| --- | --- | --- | --- | --- | --- | --- | --- | --- |
|  | Odds ratio | 95% CI of odds ratio |  | p-value | Odds ratio | 95% CI of odds ratio |  | p-value |
|  |  | Lower | Upper |  |  | Lower | Upper |  |
| Age | 1.03 | 1.01 | 1.06 | 0.008* | 1.04 | 1.01 | 1.08 | 0.009* |
| **Gender** |  |  |  |  |  |  |  |  |
| Male | Reference |  |  |  | Reference |  |  |  |
| Female | 1.7 | 1.52 | 1.9 | <0.001** | 2.08 | 1.75 | 2.46 | <0.001** |
| **Socioeconomic status** |  |  |  |  |  |  |  |  |
| Low-income | 2.15 | 1.72 | 2.7 | <0.001** | 1.49 | 1.07 | 2.1 | 0.01* |
| Middle-income | 1.48 | 1.21 | 1.82 | <0.001** | 1.01 | 0.73 | 1.38 | 0.97 |
| High-income | Reference |  |  |  | Reference |  |  |  |
| **Level of Education** |  |  |  |  |  |  |  |  |
| Pre-clinical year | Reference |  |  |  | Reference |  |  |  |
| Clinical year | 0.91 | 0.79 | 1.04 | 0.19 | 0.91 | 0.76 | 1.11 | 0.38 |
| Internship | 0.97 | 0.81 | 1.16 | 0.79 | 0.75 | 0.59 | 0.97 | 0.03* |
| **Living arrangement** |  |  |  |  |  |  |  |  |
| With family | Reference |  |  |  | Reference |  |  |  |
| With friends and colleagues | 1.77 | 1.42 | 2.20 | <0.001** | 1.31 | 1.01 | 1.71 | 0.04* |
| Alone | 1.26 | 0.91 | 1.76 | 0.16 | 1.1 | 0.72 | 1.67 | 0.65 |
| **Displacement** |  |  |  |  |  |  |  |  |
| Non-displaced | Reference |  |  |  | Reference |  |  |  |
| Displaced | 1.43 | 1.27 | 1.61 | <0.001** | 1.08 | 0.91 | 1.29 | 0.36 |
| **Living in conflict zone** |  |  |  |  |  |  |  |  |
| No | Reference |  |  |  | Reference |  |  |  |
| Yes | 1.19 | 1.02 | 1.39 | 0.02* | 1.15 | 0.94 | 1.4 | 0.15 |
| **Injury Exposure** |  |  |  |  |  |  |  |  |
| No | Reference |  |  |  | Reference |  |  |  |
| Yes | 1.34 | 1.18 | 1.51 | <0.001** | 1.02 | 0.85 | 1.21 | 0.82 |
| **Death exposure** |  |  |  |  |  |  |  |  |
| No | Reference |  |  |  | Reference |  |  |  |
| Yes | 1.28 | 1.12 | 1.46 | <0.001** | 1.5 | 1.25 | 1.79 | <0.001** |
| **Direct attacks, such as physical assaults, bombings, shootings, kidnap, or torture** |  |  |  |  |  |  |  |  |
| No | Reference |  |  |  | Reference |  |  |  |
| Yes | 1.22 | 1.05 | 1.42 | 0.007* | 1.38 | 1.14 | 1.66 | <0.001** |
| **Experiencing destruction or loss of property** |  |  |  |  |  |  |  |  |
| No | Reference |  |  |  | Reference |  |  |  |
| Yes | 1.46 | 1.12 | 1.66 | <0.001** | 1.47 | 1.25 | 1.73 | <0.001** |
| **Gender-specific violence** |  |  |  |  |  |  |  |  |
| No | Reference |  |  |  | Reference |  |  |  |
| Yes | 1.34 | 0.99 | 1.82 | 0.05 | 0.80 | 0.56 | 1.13 | 0.21 |
| **Rape or sexual violence** |  |  |  |  |  |  |  |  |
| No | Reference |  |  |  | Reference |  |  |  |
| Yes | 1.12 | 0.61 | 2.09 | 0.71 | 1.3 | 0.72 | 2.34 | 0.37 |
| **Witnessing violence against others** |  |  |  |  |  |  |  |  |
| No | Reference |  |  |  | Reference |  |  |  |
| Yes | 1.53 | 1.31 | 1.8 | <0.001** | 1.49 | 1.23 | 1.79 | <0.001** |

\*Significant at $p < 0.05$ , \*\*Significant at $p < 0.001$ .

The most significant, impactful predictive factors are the following, in descending order: low SES (OR: 2.15), living with friends and colleagues (OR: 1.77), being female (OR: 1.7), witnessing violence against others (OR: 1.53), loss or destruction of property (OR: 1.46), and suffering direct assaults (OR: 1.22).

PTSD symptoms were assessed with the ITQ questionnaire, and in total (Table 2), we found 2651 (39.9%) of the respondents fulfilled the criteria for Avoidance symptoms, 2251 (33.9%) for Reexperience, 3026 (45.6%) for the sense of current threat, 3670 (55.3) for Functional impairment, and finally, 901 (13.6%) fulfilled all four criteria for PTSD. Subsequent analysis showed significant association between PTSD and some study variables (Table 6). Gender differences were evident as females demonstrating a higher prevalence of PTSD than males. SES significantly influenced PTSD, with higher rate in low income compared to high income individuals. exposure to death, as well as exposure violence against others, were linked to higher rates of PTSD.

The logistical regression model for PTSD (Table 6) was statistically significant for ꭕ2(17) = 247.814, p<0.001, and it explained 6.7% Nagelkerke R2 of the variance, classifying 81.9% of cases, at a sensitivity of 23.2% and specificity of 91.1%, and with a positive predictive value 28.9% of and negative predictive value of 88.3%.

The most significant, impactful predictive factors are the following, in descending order: being female (OR: 2.08), exposure to death (OR: 1.5), Low SES and witnessing violence against others (OR: 1.49), destruction or loss of property (OR: 1.47),

When comparing the median (IQR) level of depression, anxiety, and insomnia (Table 1) between the respondents’ gender, females have significantly higher median score (IQR) for depression in comparison to males (12 [8–18] Vs. 9 [5–16]; p <0.001), and the same applies to anxiety (9 [5–15] vs. 7 [3–13]; p <0.001), and insomnia (7 [4–12] vs. 6 [3–11]; p < 0.001). Likewise, participants who have low income compared to middle and high income have significantly higher median scores (ITQ) for depression (13[8–19] vs.10[6–16] vs. 8[4–15]; p <0.001), at also applies to anxiety and insomnia scores.

Those living with family fared better than those who lived with friends, who in turn fared better than those who lived alone regarding depression, 11 (7-17) Vs. 12 (8-18) Vs. 13 (8-20) respectively (p<0.001), and the same applied to Anxiety, 8 (4-14) Vs. 9 (6-15) Vs. 11 (6-16) respectively (p<0.001), and insomnia, 7 (4-11) Vs. 8 (5-13) Vs. 9 (4-14) respectively (p<0.001). Those who were not displaced tended to fare better than those who were internally displaced, which in turn fared better than those externally displaced, 10 (6-16) Vs. 11 (7-17) Vs. 12 (7-18) (p<0.001) regarding depression, and 7 (3-13) Vs. 9 (5-14) Vs. 9 (5-14) (p<0.001) regarding Anxiety, and 6 (3-9) Vs. 7 (4-11) Vs. 8 (4-12) (p<0.001) regarding insomnia, respectively. The same trend goes for those in conflict zones Vs. outside of conflict zones: 12 (8-18) Vs. 11 (7-17) (p=0.005), respectively.

The relationship of these with age was a bit distinct, with all categories sharing similar levels of anxiety and giving us statistically insignificant results 8 (4-14), but those younger (<21) faring better than those older (≥21) regarding depression (10 (6-16) Vs. 11 (7-17) (p<0.001**), and regarding insomnia 7 (3-11) for <21 years old, 7 (4-11) for 21-24 years old, and 8 (4-13) for >24 years old, p<0.001.

Current level of education was also distinct in that it showed those earlier in their university education to suffer better than those who are in their clinical years and those who are interns regarding depression (11 (7-17) Vs. 11 (7-17) Vs. 11 (7-18), respectively, p=0.007*) and regarding insomnia 7 (3-11) Vs. 7 (4-11) Vs. 8 (4-13), respectively, p<0.001**), but no statistically significant difference was found regarding anxiety.

### Correlations Between Mental health parameters

A correlation analysis between mental health parameters was conducted, revealing numerous independent associations among them as shown in the (Table 7). There were statistically significant correlations between all scales in addition to significant correlations with the age of participants, level of education, number of events, current SES level, and SES changes. A higher level of depression (measured with the PHQ-9 scale) was associated with a strong positive correlation between both anxiety and insomnia symptoms (as indicated by scores on the GAD-7 and AIS-8 scales), r (6639) = 0.68-0.76, p <0.001, this also applies to the other combinations of the scores concerned. In addition, there was a significant positive weak correlation between the age of participants and their level of education, with PHQ-9, GAD-7, and AIS-8 scores, suggesting that, as the age of participants increases and as their level of education advances, there is a concomitant increase in severity of symptoms. However, there was a significant association between SES and the change to SES, and various mental health and sleep parameters, as a negative correlation exists with PHQ-9, AIS-8, and GAD-7. Indicating that a low and deteriorating income corresponds to a higher level of depression, anxiety, and insomnia symptoms.

**Table 7.**
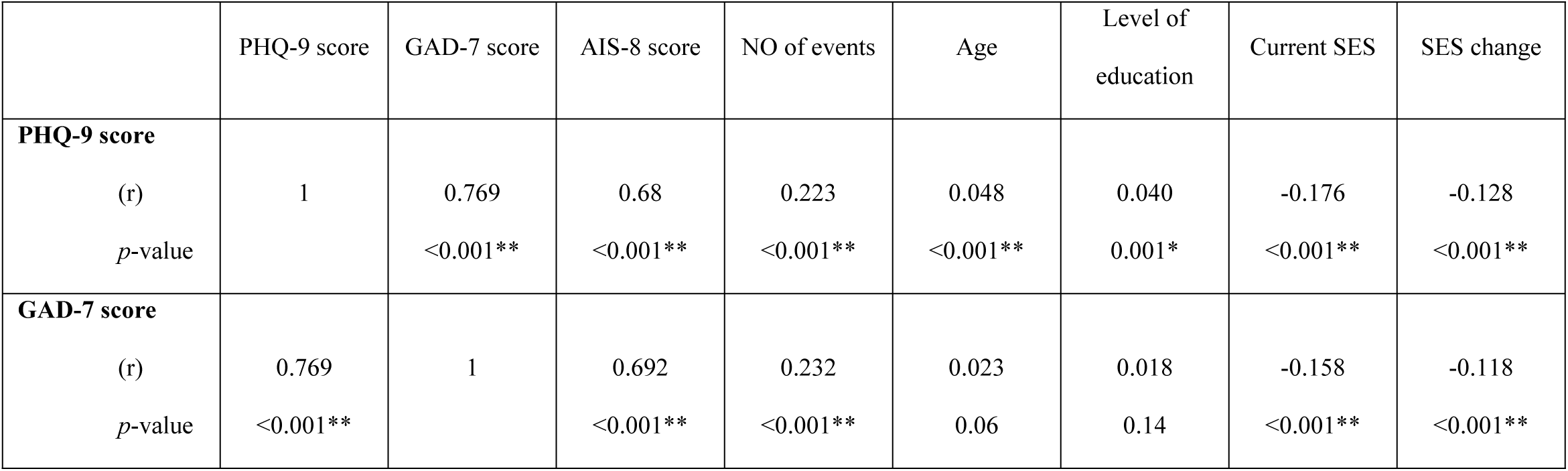
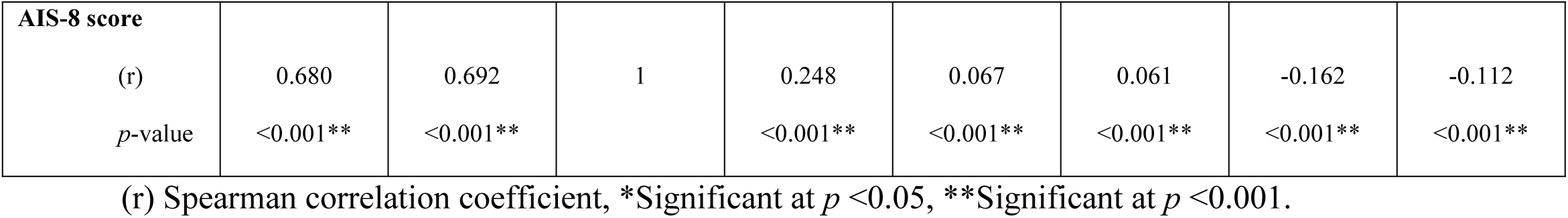
Correlations Between Mental health parameters.

## Discussion

We choose to leave our personal experience of two civil wars aside for now to maintain the objectivity of this paper. We will discuss our findings regarding the data we collected and others already available – though minimal in this specific context. We want to stress that those we reached are those on the better side of things: those with infrastructure stable enough for some 20 minutes to respond to our survey, and the means and the hope to bother at all, which effectively means that what we’ve managed to touch upon is merely the tip of the miserable, depressing iceberg of bloodshed and psychological trauma, and that those in the most desperate need were simply beyond our reach at all, despite our best efforts, and more targeted approaches must be employed in the field, hands on, by organizations with the resources to mobilize. War is war, and hell is hell, and of the two war is by far the worse, as was once said.

The purpose of this paper is to observe the prevalence of specific, morbid mental health disorders during the civil war in Sudan of April 2023. In war, it is expected to see a negative impact as the rule due to all the savagery innate to it, the bloodshed, and the upheaval, and the factors associated are myriad, of which we’d chosen a few.

We find remarkable prevalences of psychopathologies in our sample as expected during wartimes; our findings are concurrent with others considering the context for the most part, despite the deficiency regarding medical students. This study aimed to have a considerable sample size to enhance its statistical power and allow for representation for the multitudes within Sudan and their diverse backgrounds best as possible, as Sudan is a huge country in every regard, geographically, ethnically, and as a destination for those seeking education including medical students.

For depression, the findings in the recent metanalysis by Lim et al [39] of 57 studies done in 2022 show an aggregate prevalence of 28.9 % for depression Vs. our 33.7%. Anxiety had an aggregate of 30.7% Vs. our finding of 22.4%, and PTSD at 23.5% Vs. our 13.6%. other studies like that of Elhadi from the Libyan civil war [23] showed a prevalence of depression at 21.6%, both far less than in that conducted in Syria by Al Saadi at 60.6% [40], with a significant impact of financial security in that study. A study in Iraq had a prevalence more similar to the one in Syria at 52.1%, while anxiety had a prevalence of 62.5% [41].

Insomnia is a point-source for complications on both psychological and physiological levels [42], both immediately and in the future, and we find a prevalence of 11.5% of moderate and severe insomnia Vs. 49% from the recent paper from Pavlova et al. regarding Ukraine’s ongoing war [43].

These findings are to be expected considering the impact of a devastating, destabilizing civil war [44, 45]. Little attention, however, is aimed at the population of medical students during wartime despite being one of the core, most desperately needed forces to support the recovery and stabilization of a scarred society and the health of civilization both during and after wars, and despite them being a vulnerable population even during peacetime, both locally and globally [46]. Suicidal ideation had a disconcerting prevalence in our sample, with a figure that’s more severe than that found in US Veterans [47], and the general population (23%) [48], but less than the findings in Libya during their civil war at 22.7% [23]. The specific causes of such differences are a challenge to ascertain.

Regarding depression, those most severely afflicted are those aged 21-24 years old in our sample, likely reflecting their concerns regarding the upheaval in their lives both present and future, as well as their overall vulnerability as an age group, and the radical disruption to their image of themselves rooted in their plans.

Females tended to suffer the worst from all psychopathologies we’d considered as a well-known vulnerable population, we find higher prevalences across the board [49–52], as did suffering a significant SES disadvantage. These findings are well in line with others in the same vein [53–56], and Sudanese women are known to struggle with the demands of life in this developing country.

Living alone tended towards negative outcomes as expected [57–60], since living alone is a full-blown health risk where loneliness impacts negatively both the physiological and psychological health but also itself is caused by challenges to health and wellbeing. It suffices to point out that isolation is a method of torture used to break the psyche to illustrate the severity of such a casual epidemic during times of war, yet our findings were of significant AOR for Anxiety and Depression but not Insomnia and PTSD.

Being displaced seems to be a risk factor for all four conditions being considered, and our findings concur with the norm [43, 61, 62]. Being displaced results in thorough destabilization of all aspects of life and is akin to exposure, it results in vulnerabilities that may have been inconceivable before like having no access to basic facilities or infrastructure, very poor hygiene and subsequent epidemics, loss of livelihood and living standards, loss of connections and support networks, all while struggling in the face of stigma and resentment.

Material losses were also found to be a risk for all conditions considered, likely due to the reliance on what’s lost, and therefore losing all that’s dependent on what’s lost, e.g., Loss of privacy and security in the case of losing one’s home, loss of independence and flexibility, loss of status, loss of the means of production, and so on. It is no wonder that such losses are so impactful, and further insight should help alleviate the damage done by such losses, aiding in restoring health and wealth all the same, encouraging independence and production, and restoring a sense of meaning and belonging.

Living in conflict zones showed an increased AOR for Insomnia in concurrence with Pavlova’s findings [43], but we found no significant increase to depression, Anxiety, and PTSD, in contrast to findings in other studies [5, 43, 63]. This is perhaps best explained by the maintenance of social networks and the security of the familiar, preserving a sense of belonging and meaning, even under the threat of death.

Experiencing death of contacts is a well-recognized cause of depression, anxiety, PTSD, and insomnia [64–67], and our findings concur: death of contacts increased the AOR for all considered psychopathologies. These may be attributed to several consequences that include losing caretakers and providers both materially and emotionally, the loss of one’s connection to the past, to the future, and to the world, as well as being forced to face one’s mortality directly and abruptly under such distress.

Witnessing violence also resulted in an increase in negative outcomes for all four outcomes, but to lesser degree than experiencing trauma to contacts, or to the self, a relationship that caught us off guard. The same also applies to gender-specific violence, which is a well-recognized cause of trauma and impacts different identities uniquely by definition [5, 68]. Our findings regarding GSV were as expected for depression, Anxiety, and Insomnia: GSV increases the AOR. Oddly, we find no significant increase in the AOR for PTSD in contrast to other studies [69, 70]. This is concurrent with our finding that rape does not significantly increase the AOR for PTSD. After discussions regarding such findings, these are likely to be attributable to the circumstances and nature of rape and GSV, where those most severely impacted and afflicted were simply beyond our reach to begin with, meaning that this specific finding signifies a darker outcome we could not access below the waterline. These transgressions and war crimes are commonplace tactics in war, and we’d already had reports of rape, kidnap, and murder in Sudan even before the outbreak of the current war. We can only hope our line of thinking is entirely and thoroughly mistaken despite the evidence.

The level of education (Preclinical, Clinical, Internship) did not impact significantly the AOR for the considered psychopathologies in our samples.

### Limitations and author statements

This is a study provoked by our own experience during not one, but two civil wars and we have chosen the factors considered accordingly. We have seen death and got acquainted with it in savage ways, and I have personally seen men bleed to death over the course of hours with not even so much as adequate analgesia, among other, oddly less pleasant experiences of kidnap and torture, yet here we are again. We have suffered a great blow to our current and future lives, been displaced, been robbed, assaulted, kidnapped, and tortured, and we have seen our own students suffering the same as we did. Our neighbors. Our colleagues. We feel abandoned by society at large, persecuted even, accused, at times, both locally and globally. We plead for help building our livelihoods and future, to stand on our own, and thus we cry with cold, hard statistics. Our suffering is great, and it will echo throughout our lives and beyond, we only ask that we be helped get back on track while we pull ourselves from the ashes of our previous lives before all belligerence and unjustifiable deaths and loss and pain. We merely ask that we be helped to build better and do better and welcomed to do just that.

We are also acutely aware of the fact that we could not reach those who most desperately needed to be reached, those whose living conditions are so dire they have no electricity or communications, those who had suffered material losses that prevent their participation, those so depressed and traumatized they wouldn’t care to respond if even they could. we can only this helps them anyway reconnect to the world and move on with their lives, as our pleas are heard. That many are below the waterline, drowning and drowned, is a nightmare we cannot possibly stress enough, if only we could know how bad things truly are.

We would also like to note that we had originally set out to study the impact of gender as a factor, but then were faced with extreme backlash for considering such, and I quote, “Western degeneracy that no self-respecting Man (Capital M) would consider” and had subsequently had to omit such an important factor/group. We hope to repeat our study down the line, and we plan then to inquire about gender.

This study also considered especially medical students and fresh graduates, and perhaps our findings apply to similar groups, however, that is beyond our scope.

## Conclusion

War is strongly condemned in the modern world, and its impacts are myriad and thoroughly negative bar the most unusual views. We find a great deal of suffering in our population, and our study highlights the fact by observing the prevalence of four psychological disorders in the population of medical students and fresh medical graduates who suffered the war in Sudan. The negative aspects are reasonable and consistent for the most part with what’s to be expected: losing one’s place in society and society itself, losing one’s contacts and property, and being injured physically and transgressed against are all harmful with lasting effects on the persons involved, and observing such results would be instrumental for effective management of sequalae.

## Recommendations

Our recommendations regarding our findings are in line with those of the UN, and we find it reasonable considering our findings: those left without an established life in the present and in the future are prone to manifestations of psychological distress including depression, anxiety, PTSD, and insomnia.

To boot, that the globe has such a need for doctors, current and future, everywhere, in every specialty, yet so willingly discards such huge numbers teeters towards unfathomable cruelty to all stakeholders and totters towards being a poor punchline to a joke that’s all a punch to the gut, though rarely it makes contact as it did during the years of COVID-19.

What we plead for is a way for the survivors to reestablish their lives and carry on, and having a career is an essential aspect of that, a career that starts with a completed education, conserves dignity, permits integration, facilitates ascendancy, and inspires sublimation. Many of us survivors can go on with minimal aid: simply provide the examination centers and be willing to help us reintegrate into the medical pipeline, especially those on the verge of graduation/recent graduates as they suffer the most profound losses on par with mutilation, require the least support: are you on par with others as you claim? and are the most rewarding: we now have another desperately needed practitioner! The available research supports that point beyond doubt, and yet here we are, having to go through hundreds of thousands for a sample of a few thousands to state the obvious, a dead horse that for some reason we are still savagely and relentlessly flogging…

And for those who welcomed our efforts and rushed to our aid; we owe you a debt of gratitude long as we may live as physicians doing what we can, where we can.

## Data Availability

The raw data generated or analyzed during the course of this research is available in online repositories (openICPSR) http://doi.org/10.3886/E198524V2

## Acknowledgment

We would like to express our gratitude to all those who contributed to this work.

## Supporting information

S1 Questionnaire English: English version of questionnaire.

S2 Questionnaire Arabic: Arabic version of questionnaire.

S3 Table 1: Frequencies and percentages of exposure to events.

S4 Table2: Complete list of Members of the SudaMedReCollab study group.

S5 STROBE Checklist: STROBE Statement Checklist of cross-sectional studies

## References

1. Khalid Abdelaziz NE. Sudan clashes kill at least 25 in power struggle between army, paramilitaries: Reuters; 2023 [cited 2023]. Available from: https://www.reuters.com/world/africa/heavy-gunfire-heard-south-sudanese-capital-khartoum-witnesses-2023-04-15/.

2. Migration IOf. DTM Sudan’s Internally Displaced Persons 2023 Estimates | Displacement Tracking Matrix 2024. Available from: https://dtm.iom.int/reports/dtm-sudans-internally-displaced-persons-2023-estimates.

3. Elshazali OH, Abdullahi H, Karrar ZA. Progress, challenges and partnerships of teaching medical professionalism in medical schools in Sudan: the success story of Sudan Medical Council. Sudan J Paediatr. 2021;21(2):110–5. Epub 2021/01/01. doi: 10.24911/sjp.106-1622725530. PubMed PMID: 35221421; PubMed Central PMCID: PMCPMC8879352.

4. Sudan Country Profile 2023. Available from: https://www.bbc.com/news/world-africa-14094995.

5. Murthy RS, Lakshminarayana R. Mental health consequences of war: a brief review of research findings. World Psychiatry. 2006;5(1):25–30. PubMed PMID: 16757987; PubMed Central PMCID: PMCPMC1472271.

6. Cardozo BL, Bilukha OO, Crawford CA, Shaikh I, Wolfe MI, Gerber ML, et al. Mental health, social functioning, and disability in postwar Afghanistan. JAMA. 2004;292(5):575–84. doi: 10.1001/jama.292.5.575. PubMed PMID: 15292083.

7. Scholte WF, Olff M, Ventevogel P, de Vries GJ, Jansveld E, Cardozo BL, et al. Mental health symptoms following war and repression in eastern Afghanistan. JAMA. 2004;292(5):585–93. doi: 10.1001/jama.292.5.585. PubMed PMID: 15292084.

8. Di Giovanni J. Madness visible: a memoir of war: Vintage; 2007.

9. Rose M. Fighting for peace, Bosnia 1994: Vintage; 1998.

10. Beloff N. Yugoslavia: an avoidable war: New European Publications; 1997.

11. Westermeyer J. Health of Albanians and Serbians following the war in Kosovo: studying the survivors of both sides of armed conflict. JAMA. 2000;284(5):615–6. doi: 10.1001/jama.284.5.615. PubMed PMID: 10918709.

12. Lopes Cardozo B, Vergara A, Agani F, Gotway CA. Mental health, social functioning, and attitudes of Kosovar Albanians following the war in Kosovo. JAMA. 2000;284(5):569–77. doi: 10.1001/jama.284.5.569. PubMed PMID: 10918702.

13. Salama P, Spiegel P, Van Dyke M, Phelps L, Wilkinson C. Mental health and nutritional status among the adult Serbian minority in Kosovo. JAMA. 2000;284(5):578–84. doi: 10.1001/jama.284.5.578. PubMed PMID: 10918703.

14. Boehnlein JK, Kinzie JD, Sekiya U, Riley C, Pou K, Rosborough B. A ten-year treatment outcome study of traumatized Cambodian refugees. J Nerv Ment Dis. 2004;192(10):658–63. doi: 10.1097/01.nmd.0000142033.79043.9d. PubMed PMID: 15457108.

15. Mollica RF, Donelan K, Tor S, Lavelle J, Elias C, Frankel M, et al. The effect of trauma and confinement on functional health and mental health status of Cambodians living in Thailand-Cambodia border camps. JAMA. 1993;270(5):581–6. PubMed PMID: 8331755.

16. Ahmad A, Sofi MA, Sundelin-Wahlsten V, von Knorring AL. Posttraumatic stress disorder in children after the military operation “Anfal” in Iraqi Kurdistan. Eur Child Adolesc Psychiatry. 2000;9(4):235–43. doi: 10.1007/s007870070026. PubMed PMID: 11202098.

17. Karam EG, Howard DB, Karam AN, Ashkar A, Shaaya M, Melhem N, et al. Major depression and external stressors: the Lebanon Wars. Eur Arch Psychiatry Clin Neurosci. 1998;248(5):225–30. doi: 10.1007/s004060050042. PubMed PMID: 9840368.

18. Kinzie JD, Sack W, Angell R, Clarke G, Ben R. A three-year follow-up of Cambodian young people traumatized as children. J Am Acad Child Adolesc Psychiatry. 1989;28(4):501–4. doi: 10.1097/00004583-198907000-00006. PubMed PMID: 2768143.

19. Paardekooper B, de Jong JT, Hermanns JM. The psychological impact of war and the refugee situation on South Sudanese children in refugee camps in Northern Uganda: an exploratory study. J Child Psychol Psychiatry. 1999;40(4):529–36. PubMed PMID: 10357160.

20. Al-Krenawi A, Graham JR, Sehwail MA. Mental Health and Violence/Trauma in Palestine: Implications for Helping Professional Practice. Journal of Comparative Family Studies. 2004;35(2):185–209.

21. de Jong JT, Komproe IH, Van Ommeren M, El Masri M, Araya M, Khaled N, et al. Lifetime events and posttraumatic stress disorder in 4 postconflict settings. JAMA. 2001;286(5):555–62. doi: 10.1001/jama.286.5.555. PubMed PMID: 11476657.

22. Kurapov A, Danyliuk I, Loboda A, Kalaitzaki A, Kowatsch T, Klimash T, et al. Six months into the war: a first-wave study of stress, anxiety, and depression among in Ukraine. Frontiers in Psychiatry. 2023;14. doi: 10.3389/fpsyt.2023.1190465.

23. Elhadi M, Buzreg A, Bouhuwaish A, Khaled A, Alhadi A, Msherghi A, et al. Psychological Impact of the Civil War and COVID-19 on Libyan Medical Students: A Cross-Sectional Study. Front Psychol. 2020;11:570435. Epub 20201026. doi: 10.3389/fpsyg.2020.570435. PubMed PMID: 33192858; PubMed Central PMCID: PMCPMC7649391.

24. Mohamed OGN, Mohamed EGN, Ahmed R, Aburas L, Ali M, Hamdan HZ. Depression, Anxiety, and Stress among Sudanese Medical Students during the COVID-19 Lockdown Period. Open Access Macedonian Journal of Medical Sciences. 2022;10(B):1365–71. doi: 10.3889/oamjms.2022.9432.

25. Levis B, Benedetti A, Thombs BD. Accuracy of Patient Health Questionnaire-9 (PHQ-9) for screening to detect major depression: individual participant data meta-analysis. Bmj. 2019;365:l1476. Epub 2019/04/11. doi: 10.1136/bmj.l1476. PubMed PMID: 30967483; PubMed Central PMCID: PMCPMC6454318 at www.icmje.org/coi_disclosure.pdf (available on request from the corresponding author)

26. AlHadi AN, AlAteeq DA, Al-Sharif E, Bawazeer HM, Alanazi H, AlShomrani AT, et al. An arabic translation, reliability, and validation of Patient Health Questionnaire in a Saudi sample. Ann Gen Psychiatry. 2017;16:32. Epub 2017/09/08. doi: 10.1186/s12991-017-0155-1. PubMed PMID: 28878812; PubMed Central PMCID: PMCPMC5585978.

27. Kroenke K, Spitzer RL, Williams JB. The PHQ-9: validity of a brief depression severity measure. J Gen Intern Med. 2001;16(9):606–13. Epub 2001/09/15. doi: 10.1046/j.1525-1497.2001.016009606.x. PubMed PMID: 11556941; PubMed Central PMCID: PMCPMC1495268.

28. Urtasun M, Daray FM, Teti GL, Coppolillo F, Herlax G, Saba G, et al. Validation and calibration of the patient health questionnaire (PHQ-9) in Argentina. BMC Psychiatry. 2019;19(1):291. Epub 2019/09/20. doi: 10.1186/s12888-019-2262-9. PubMed PMID: 31533674; PubMed Central PMCID: PMCPMC6751851.

29. Cloitre M, Shevlin M, Brewin CR, Bisson JI, Roberts NP, Maercker A, et al. The International Trauma Questionnaire: development of a self-report measure of ICD-11 PTSD and complex PTSD. Acta Psychiatr Scand. 2018;138(6):536–46. Epub 2018/09/05. doi: 10.1111/acps.12956. PubMed PMID: 30178492.

30. Hyland P, Shevlin M, Brewin CR, Cloitre M, Downes AJ, Jumbe S, et al. Validation of post-traumatic stress disorder (PTSD) and complex PTSD using the International Trauma Questionnaire. Acta Psychiatr Scand. 2017;136(3):313–22. Epub 2017/07/12. doi: 10.1111/acps.12771. PubMed PMID: 28696531.

31. Vallières F, Ceannt R, Daccache F, Abou Daher R, Sleiman J, Gilmore B, et al. ICD-11 PTSD and complex PTSD amongst Syrian refugees in Lebanon: the factor structure and the clinical utility of the International Trauma Questionnaire. Acta Psychiatrica Scandinavica. 2018;138(6):547–57. doi: 10.1111/acps.12973.

32. Spitzer RL, Kroenke K, Williams JBW, Löwe B. A Brief Measure for Assessing Generalized Anxiety Disorder: The GAD-7. Archives of Internal Medicine. 2006;166(10):1092–7. doi: 10.1001/archinte.166.10.1092.

33. Löwe B, Decker O, Müller S, Brähler E, Schellberg D, Herzog W, et al. Validation and standardization of the Generalized Anxiety Disorder Screener (GAD-7) in the general population. Med Care. 2008;46(3):266–74. Epub 2008/04/05. doi: 10.1097/MLR.0b013e318160d093. PubMed PMID: 18388841.

34. Ruiz MA, Zamorano E, García-Campayo J, Pardo A, Freire O, Rejas J. Validity of the GAD-7 scale as an outcome measure of disability in patients with generalized anxiety disorders in primary care. J Affect Disord. 2011;128(3):277–86. Epub 2010/08/10. doi: 10.1016/j.jad.2010.07.010. PubMed PMID: 20692043.

35. Kroenke K, Spitzer RL, Williams JB, Monahan PO, Löwe B. Anxiety disorders in primary care: prevalence, impairment, comorbidity, and detection. Ann Intern Med. 2007;146(5):317–25. Epub 2007/03/07. doi: 10.7326/0003-4819-146-5-200703060-00004. PubMed PMID: 17339617.

36. Soldatos CR, Dikeos DG, Paparrigopoulos TJ. Athens Insomnia Scale: validation of an instrument based on ICD-10 criteria. J Psychosom Res. 2000;48(6):555–60. Epub 2000/10/18. doi: 10.1016/s0022-3999(00)00095-7. PubMed PMID: 11033374.

37. Hallit S, Haddad C, Hallit R, Al Karaki G, Malaeb D, Sacre H, et al. Validation of selected sleeping disorders related scales in Arabic among the Lebanese Population. Sleep and Biological Rhythms. 2019;17(2):183–9. doi: 10.1007/s41105-018-0196-0.

38. Soldatos CR, Dikeos DG, Paparrigopoulos TJ. The diagnostic validity of the Athens Insomnia Scale. J Psychosom Res. 2003;55(3):263–7. Epub 2003/08/23. doi: 10.1016/s0022-3999(02)00604-9. PubMed PMID: 12932801.

39. Lim I, Tam WWS, Chudzicka-Czupala A, McIntyre RS, Teopiz KM, Ho RC, et al. Prevalence of depression, anxiety and post-traumatic stress in war- and conflict-afflicted areas: A meta-analysis. Front Psychiatry. 2022;13:978703. Epub 20220916. doi: 10.3389/fpsyt.2022.978703. PubMed PMID: 36186881; PubMed Central PMCID: PMCPMC9524230.

40. Al Saadi T, Zaher Addeen S, Turk T, Abbas F, Alkhatib M. Psychological distress among medical students in conflicts: a cross-sectional study from Syria. BMC Med Educ. 2017;17(1):173. Epub 20170920. doi: 10.1186/s12909-017-1012-2. PubMed PMID: 28931387; PubMed Central PMCID: PMCPMC5607487.

41. Rasheed AG, Hussein AG. Depression, anxiety, and stress among medical students of College of Medicine, Hawler Medical University, Erbil, Iraq. Zanco Journal of Medical Sciences (Zanco J Med Sci). 2019;23(2):143–52.

42. Fernandez-Mendoza J, Vgontzas AN. Insomnia and its impact on physical and mental health. Curr Psychiatry Rep. 2013;15(12):418. doi: 10.1007/s11920-013-0418-8. PubMed PMID: 24189774; PubMed Central PMCID: PMCPMC3972485.

43. Pavlova I, Rogowska AM. Exposure to war, war nightmares, insomnia, and war-related posttraumatic stress disorder: A network analysis among university students during the war in Ukraine. Journal of Affective Disorders. 2023;342:148–56. doi: 10.1016/j.jad.2023.09.003.

44. Steel Z, Chey T, Silove D, Marnane C, Bryant RA, van Ommeren M. Association of torture and other potentially traumatic events with mental health outcomes among populations exposed to mass conflict and displacement: a systematic review and meta-analysis. JAMA. 2009;302(5):537–49. doi: 10.1001/jama.2009.1132. PubMed PMID: 19654388.

45. Wells TS, Miller SC, Adler AB, Engel CC, Smith TC, Fairbank JA. Mental health impact of the Iraq and Afghanistan conflicts: a review of US research, service provision, and programmatic responses. Int Rev Psychiatry. 2011;23(2):144–52. doi: 10.3109/09540261.2011.558833. PubMed PMID: 21521083.

46. Maser B, Danilewitz M, Guerin E, Findlay L, Frank E. Medical Student Psychological Distress and Mental Illness Relative to the General Population: A Canadian Cross-Sectional Survey. Acad Med. 2019;94(11):1781–91. doi: 10.1097/ACM.0000000000002958. PubMed PMID: 31436626.

47. Nichter B, Stein MB, Norman SB, Hill ML, Straus E, Haller M, et al. Prevalence, Correlates, and Treatment of Suicidal Behavior in US Military Veterans: Results From the 2019-2020 National Health and Resilience in Veterans Study. J Clin Psychiatry. 2021;82(5). Epub 20210810. doi: 10.4088/JCP.20m13714. PubMed PMID: 34383391.

48. Nouri E, Moradi Y, Moradi G. The global prevalence of suicidal ideation and suicide attempts among men who have sex with men: a systematic review and meta-analysis. European Journal of Medical Research. 2023;28(1):361. doi: 10.1186/s40001-023-01338-6.

49. Weissman MM, Klerman GL. Sex differences and the epidemiology of depression. Arch Gen Psychiatry. 1977;34(1):98–111. doi: 10.1001/archpsyc.1977.01770130100011. PubMed PMID: 319772.

50. Tian-Ci Quek T, Wai-San Tam W, X. Tran B, Zhang M, Zhang Z, Su-Hui Ho C, et al. The Global Prevalence of Anxiety Among Medical Students: A Meta-Analysis. International Journal of Environmental Research and Public Health [Internet]. 2019; 16(15).

51. Ng LC, Stevenson A, Kalapurakkel SS, Hanlon C, Seedat S, Harerimana B, et al. National and regional prevalence of posttraumatic stress disorder in sub-Saharan Africa: A systematic review and meta-analysis. PLOS Medicine. 2020;17(5):e1003090. doi: 10.1371/journal.pmed.1003090.

52. Zhang B, Wing Y-K. Sex differences in insomnia: a meta-analysis. Sleep. 2006;29(1):85–93.

53. Lorant V, Deliège D, Eaton W, Robert A, Philippot P, Ansseau M. Socioeconomic Inequalities in Depression: A Meta-Analysis. American Journal of Epidemiology. 2003;157(2):98–112. doi: 10.1093/aje/kwf182.

54. Barbek RME, Makowski AC, von dem Knesebeck O. Social inequalities in health anxiety: A systematic review and meta-analysis. Journal of Psychosomatic Research. 2022;153:110706. doi: 10.1016/j.jpsychores.2021.110706.

55. Sosso FAE, Matos E, Papadopoulos D. Social disparities in sleep health of African populations: A systematic review and meta-analysis of observational studies. Sleep Health. 2023;9(6):828–45. doi: 10.1016/j.sleh.2023.08.021.

56. Brewin CR, Andrews B, Valentine JD. Meta-analysis of risk factors for posttraumatic stress disorder in trauma-exposed adults. Journal of consulting and clinical psychology. 2000;68(5):748.

57. Wu D, Liu F, Huang S. Assessment of the relationship between living alone and the risk of depression based on longitudinal studies: A systematic review and meta-analysis. Front Psychiatry. 2022;13:954857. Epub 20220830. doi: 10.3389/fpsyt.2022.954857. PubMed PMID: 36111305; PubMed Central PMCID: PMCPMC9468273.

58. Jacob L, Haro JM, Koyanagi A. Relationship between living alone and common mental disorders in the 1993, 2000 and 2007 National Psychiatric Morbidity Surveys. PLOS ONE. 2019;14(5):e0215182. doi: 10.1371/journal.pone.0215182.

59. Morina N, Stam K, Pollet TV, Priebe S. Prevalence of depression and posttraumatic stress disorder in adult civilian survivors of war who stay in war-afflicted regions. A systematic review and meta-analysis of epidemiological studies. Journal of Affective Disorders. 2018;239:328–38. doi: 10.1016/j.jad.2018.07.027.

60. Hom MA, Chu C, Rogers ML, Joiner TE. A Meta-Analysis of the Relationship Between Sleep Problems and Loneliness. Clinical Psychological Science. 2020;8(5):799–824. doi: 10.1177/2167702620922969.

61. Bedaso A, Duko B. Epidemiology of depression among displaced people: A systematic review and meta-analysis. Psychiatry Res. 2022;311:114493. Epub 20220308. doi: 10.1016/j.psychres.2022.114493. PubMed PMID: 35316692.

62. Oakley LD, Kuo W-c, Kowalkowski JA, Park W. Meta-Analysis of Cultural Influences in Trauma Exposure and PTSD Prevalence Rates. Journal of Transcultural Nursing. 2021;32(4):412–24. doi: 10.1177/1043659621993909. PubMed PMID: 33593236.

63. Carpiniello B. The Mental Health Costs of Armed Conflicts-A Review of Systematic Reviews Conducted on Refugees, Asylum-Seekers and People Living in War Zones. Int J Environ Res Public Health. 2023;20(4). Epub 20230206. doi: 10.3390/ijerph20042840. PubMed PMID: 36833537; PubMed Central PMCID: PMCPMC9957523.

64. Breslau N, Kessler RC, Chilcoat HD, Schultz LR, Davis GC, Andreski P. Trauma and posttraumatic stress disorder in the community: the 1996 Detroit Area Survey of Trauma. Arch Gen Psychiatry. 1998;55(7):626–32. doi: 10.1001/archpsyc.55.7.626. PubMed PMID: 9672053.

65. Kessler RC, Sonnega A, Bromet E, Hughes M, Nelson CB. Posttraumatic stress disorder in the National Comorbidity Survey. Arch Gen Psychiatry. 1995;52(12):1048–60. doi: 10.1001/archpsyc.1995.03950240066012. PubMed PMID: 7492257.

66. Keyes KM, Pratt C, Galea S, McLaughlin KA, Koenen KC, Shear MK. The burden of loss: unexpected death of a loved one and psychiatric disorders across the life course in a national study. Am J Psychiatry. 2014;171(8):864–71. doi: 10.1176/appi.ajp.2014.13081132. PubMed PMID: 24832609; PubMed Central PMCID: PMCPMC4119479.

67. Monk TH, Germain A, Reynolds CF. Sleep Disturbance in Bereavement. Psychiatr Ann. 2008;38(10):671–5. doi: 10.3928/00485713-20081001-06. PubMed PMID: 20179790; PubMed Central PMCID: PMCPMC2826218.

68. Kastrup MC. Mental health consequences of war: gender specific issues. World Psychiatry. 2006;5(1):33–4. PubMed PMID: 16757990; PubMed Central PMCID: PMCPMC1472268.

69. Chivers-Wilson KA. Sexual assault and posttraumatic stress disorder: a review of the biological, psychological and sociological factors and treatments. Mcgill J Med. 2006;9(2):111–8. PubMed PMID: 18523613; PubMed Central PMCID: PMCPMC2323517.

70. Tenaw LA, Aragie MW, Ayele AD, Kokeb T, Yimer NB. Medical and psychological consequences of rape among survivors during armed conflicts in northeast Ethiopia. PLoS One. 2022;17(12):e0278859. Epub 20221212. doi: 10.1371/journal.pone.0278859. PubMed PMID: 36508404; PubMed Central PMCID: PMCPMC9744300.

